# Why women choose abortion through telemedicine outside the formal health sector in Germany? A mixed-methods study

**DOI:** 10.1101/2020.09.08.20190249

**Authors:** Kristina Killinger, Sophie Günther, Hazal Atay, Rebecca Gomperts, Margit Endler

## Abstract

**Introduction:** In April 2019 the abortion telemedicine service Women on Web (WoW) opened their helpdesk to Germany and saw a progressive rise in consultations. Our aim was to understand the motivations, and perceived barriers to access, for women who choose telemedicine abortion outside the formal health sector in Germany.

**Methods:** We conducted a parallel convergent mixed-methods study among 1090 women in Germany, who requested medical abortion through WoW between January 1^st^ and December 31^st^, 2019. We performed a cross-sectional study of data contained in online consultations and a content analysis of 108 email texts. Analysis was done until saturation; results were merged, and triangulation was used to validate results.

**Results:** Frequent reported reasons for choosing telemedicine abortion in the consultation forms were *“I need to keep the abortion a secret from my partner or family”* (48%) and *“I would rather keep my abortion private”* (48%). The content analysis developed two main themes and seven subsidiary categories: 1) internal motivations for seeking telemedicine abortion encompassing i) autonomy, ii) perception of external threat, iii) shame and stigma, and 2) external barriers to formal abortion care, encompassing: (iv) financial stress, v) logistic barriers to access vi) provider attitudes, and vii) vulnerability of foreigners). The findings in the quantitative and qualitative analysis were consistent.

**Conclusion:** Women in Germany who choose telemedicine abortion outside the formal health sector do so both from a place of empowerment and a place of disempowerment. Numerous barriers to abortion access exist in the formal sector which are of special relevance to vulnerable groups such as adolescents and undocumented immigrants.

**Key message points:**

- When Women on Web, an abortion telemedicine service operating outside the formal health care sector, opened in Germany in April 2019, 1205 women consulted the service in the first nine months.
- Women who choose telemedicine abortion do so both from a position of empowerment, for reasons of autonomy, and from a position of disempowerment and lack of autonomy.
- Numerous barriers to abortion access, as permitted by German law exist in the formal health sector, which may most impact vulnerable groups such as adolescents, women with low financial means, and undocumented immigrants.

## Introduction

In April 2019 Women on Web (WoW), an abortion telemedicine service that usually serves women in countries where abortion is legally restricted, opened their helpdesk to women in Germany. Between April and December 2019, consultations increased from 44 to 193 per month, suggesting that there existed a demand for abortion services that was not being met in the formal health sector. The abortion consultation process through WoW has been described previously (1, 2). Based on available data, abortion by telemedicine has similar clinical outcomes to in-person abortion, and women chose telemedicine abortion for a multitude of reasons (3) (4).

In Germany elective abortion is technically illegal but formally allowed through a broad legal clause that permits abortion without prosecution up to 12 weeks’ gestation when it is done through a “consultation process” entailing an appointment with a state-approved agency, three days of reflection, and a physician-administered abortion (5). The advertisement of abortion services however can technically be penalised with imprisonment, and conscientious objection to abortion, the possibility of by law refusing to provide abortion care for personal beliefs, is permitted (5). Abortion providers are asked to register with the General Medical Council but information about who and where these doctors are is not accessible to the public (6) (7) In 2018, 100 986 abortions were performed in Germany, of which 96.2% occurred through the consultation process (8).

To our knowledge, there are no published studies on why women in Germany opt for abortion outside the formal health sector. Our aim was to understand the motivations, and perceived barriers to access, for women in Germany choosing telemedicine abortion over formal services.

## Methods

We performed a convergent parallel mixed methods study consisting of a cross-sectional study and a content analysis. This methodology means that both data components were collected simultaneously, analysed independently but accorded equal weight, and merged for interpretation. We triangulated the quantitative and qualitative data to validate results.

### Cross-sectional analysis

We retrieved anonymized data from consultations sent from Germany, in German or English, to WoW between January 1^st^ and December 31^st^, 2019.

We excluded consultations that were duplicate, not in English or German, from US military bases, or which had been filled out by an intermediary for women not residing in Germany. We summarized participants background characteristics and evaluated associations between age categories (with adolescents defined as women aged ≤18 years), population size of town of residence, immigration status, and categorical reasons for choosing abortion through WoW.

Continuous data were summarized as means and standard deviation or median and interquartile range according to the data’s distribution. Categorical data were summarized as frequencies. Associations between socio-demographic characteristics and reasons for choosing online abortion were tested using Pearson’s Chi-square test or Fisher’s exact test. Effect size was expressed as odds ratios (OR) with 95% confidence intervals (CI). Data analysis was performed using Stata version 16.0 (StataCorp. 2016).

### Content analysis

After submitting an online consultation to WoW, women in Germany receive an email telling them that abortion is available in the formal health care sector followed by this question:

*“If you feel you are unable to access abortion services in Germany, can you please tell us a bit more about why. The doctor will review your request for help, and we will let you know as soon as possible if we can help you in any way (2).”*

We performed a content analysis of the emails sent in response to this question. Two researchers (KK; SG) simultaneously identified the main themes and recurring categories in the text starting from January and December 2019 respectively and working consecutively until saturation after which a further 28 emails were analysed from mid-year 2019 to confirm saturation.

The complete text was then reread and discussed with a third researcher (ME) for consistency and accuracy. We used systematic coding to categorize and derive subcategories. We analysed the text at the manifest level meaning that the we used the apparent meaning, as opposed to the “latent” or underlying meaning, of the text. Units of information were analysed separately and then case-based to evaluate women’s primary and secondary motivations. We quantified recurring categories to contextualise the findings with the quantitative results.

### Data validation

We used methodological and researcher triangulation to mitigate researcher bias and increase reliability. The quantitative and qualitative outcomes were interpreted jointly and contrasted to nuance findings and identify contradictions.

Public-patient involvement was not engaged in the study design. The research was approved by the Ethics Committee at Karolinska Institutet, Sweden (Dnr 2009/2072-31/2).

## RESULTS

### Cross-sectional study

Between January1^st^ and December 31^st^ 2019, WoW received 1208 consultations from Germany of which 1090 were included after exclusions (Figure1).

**Figure 1:**
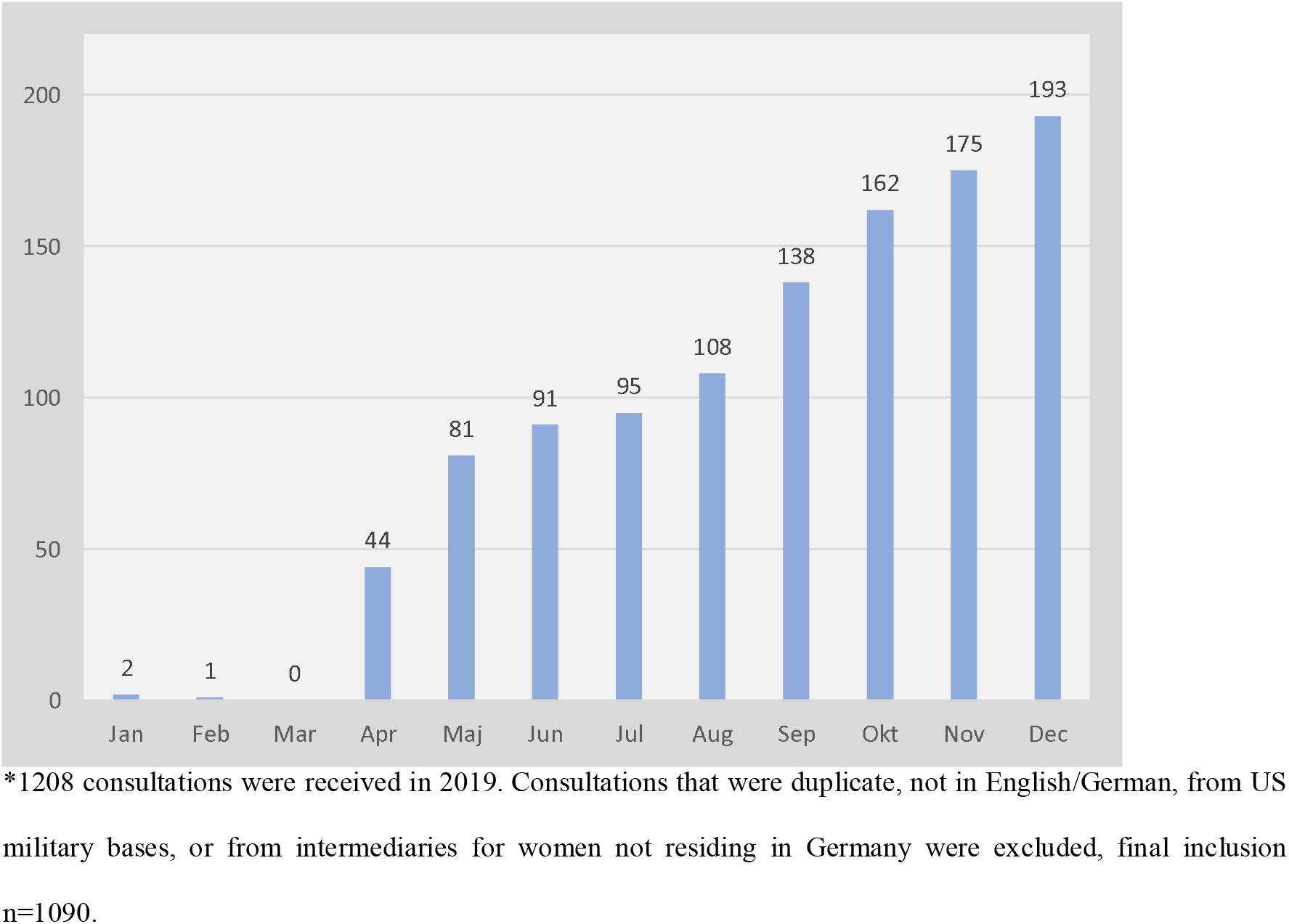
Number of consultations for medical abortion received by Women on Web per month from Germany in 2019 (n = 1090)

The median age among participants was 29 years (+/− 9 years). Median gestational age was 5 weeks(w) 6 days(d), ranging from 3w0d to 18w1d. Background characteristics of participants, categorical reasons for requesting telemedicine abortion, and associations to background variables are shown in Tables 1,2 and Supplementary Table 1 respectively.

**Table 1:** Background and pregnancy-related characteristics of 109 women in Germany requesting an abortion through Women on Web between January 1^st^ and December 31^st^ 2019

| *Continuous variables* | *Median (IQR, range)* |
| --- | --- |
| **Gestational age in days<sup>a</sup>** (missing=189) | 36 (11, 21-127) |
| **Age in years** | 29 (9; 12-53) |
| *Categorical variables* | *% (n)* |
| **Age in years** |  |
| ≤18 | 6.4 (70) |
| 19-29 | 49.3 (537) |
| 30-40 | 40 (436) |
| ≥41 | 4.3 (47) |
| **Population size of town of residence<sup>b</sup>** |  |
| Population ≥100.000 | 35.8 (390) |
| Population <100.000 | 64.2 (700) |
| **Undocumented immigrant** | 5.0 (54) |
| **Gravidity<sup>b</sup>** (missing=48) |  |
| 0 | 17.9 (186) |
| 1 | 26.1 (272) |
| 2 | 20.7 (216) |
| 3 | 18.4 (192) |
| >3 | 16.9 (176) |
| **Parity** (missing=55) |  |
| 0 | 45.3 (469) |
| 1 | 23.2 (240) |
| 2 | 20.2 (209) |
| >2 | 11.3 (117) |
| **Previous abortions** (missing=88) |  |
| 0 | 75.3 (754) |
| 1 | 19.1 (191) |
| >1 | 5.7 (57) |
| **Previous caesarean section** (missing=1) | 21.5 (234) |
| **Reasons for unwanted pregnancy** (missing=6) |  |
| No use of contraceptives | 29.7 (322) |
| Contraceptives failed | 64.3 (697) |
| Rape | 5.7 (62) |
| **Reasons for abortion<sup>c</sup>** (missing=9) |  |
| Cannot have child at this point of life | 58.9 (637) |
| Financial situation | 46.6 (504) |
| Family is complete | 26.5 (286) |
| Age (too young/old) | 26.1 (282) |
| Want to finish school | 24.5 (265) |
| Illness | 1.9 (21) |

*a Values for days since last menstrual period (LMP) <21 days re-coded as missing data since as a pregnancy test rarely detects U-HCG before this time.*
*b Questions not regarding medical eligibility are optional which explains missing data..*
*c Multiple responses allowed, total response percentages exceed 100%.*

**Table 2.**
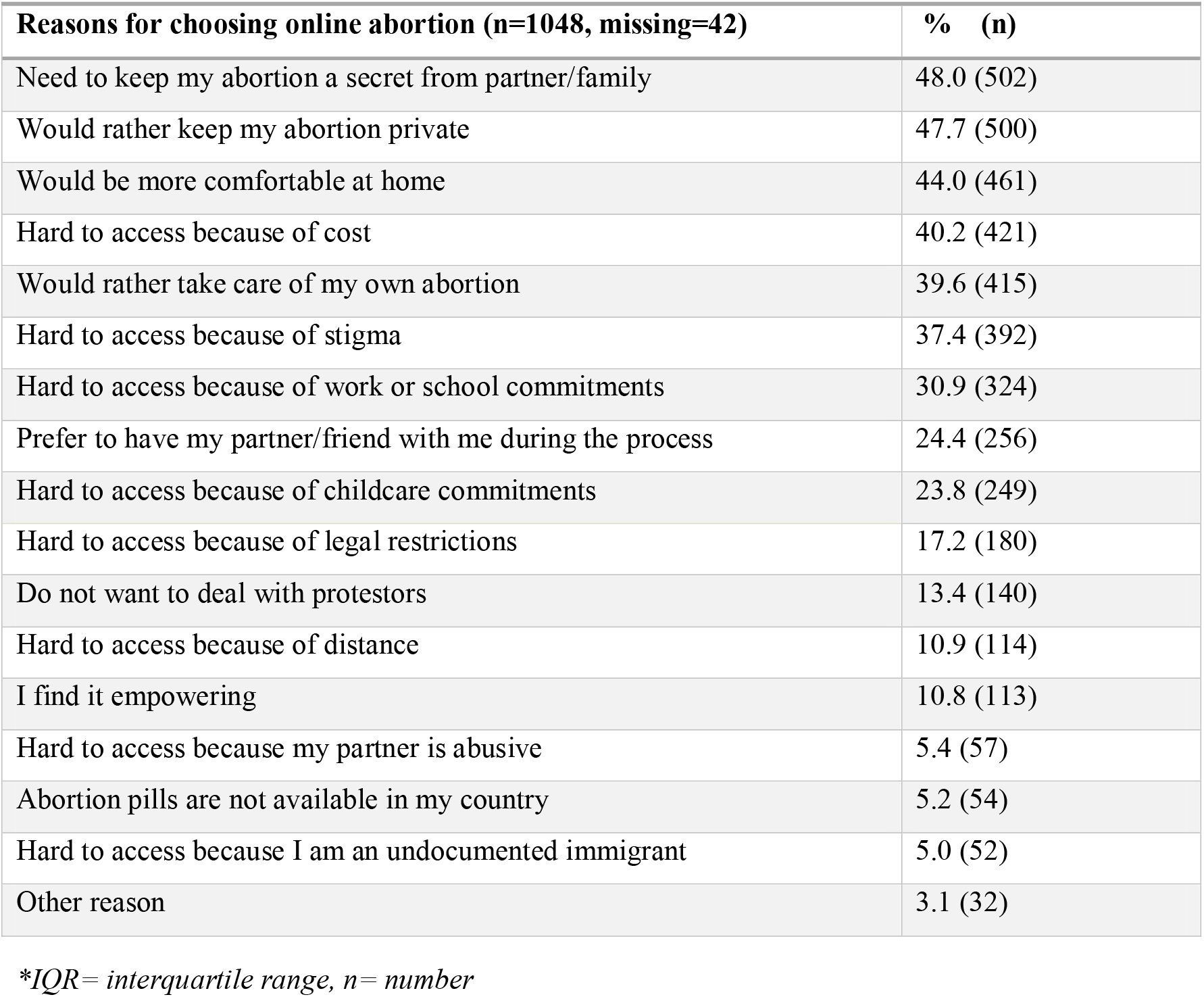
Categorical reasons for choosing online abortion over formal abortion services amongwomen in Germany requesting abortion from Women on Web January 1^st^ to December 31^st^

### Content analysis

WoW received 255 emails following online consultations from Germany during 2019. We assessed that saturation was achieved after the analysis of 80 emails, we included a further 28 emails to validate this. In total we analysed 108 emails which varied in length from 10 to 400 words.

We developed two main themes and seven categories subsidiary to these themes: 1) internal motivations for seeking telemedicine abortion encompassing i) autonomy, ii) perception of external threat, and iii) shame/fear of stigma, and 2) external barriers to formal abortion care encompassing;: iv) financial stress, v) logistic barriers to access, vi) provider attitudes, and vii) the vulnerability of foreigners. A scheme of themes, categories and subcategories is shown in figure 2.

**Figure 2:**
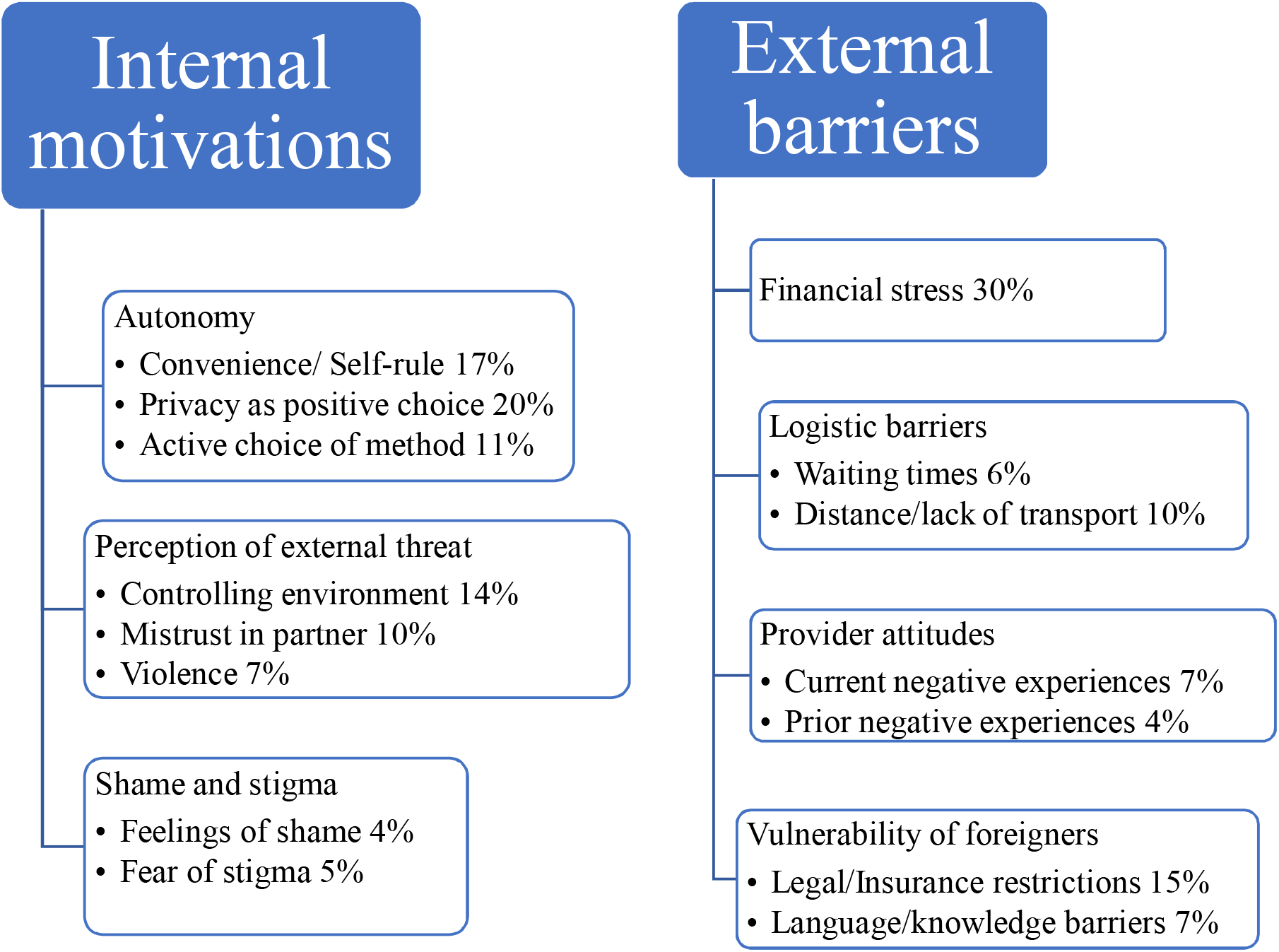
Schematic presentation of derived themes, categories and subcategories with corresponding frequencies from 108 emails sent to Women on Web between January 1^st^ and December 31^st^, 2019 explaining their need of an online abortion

### Integrated results

#### Autonomy

In the cross-sectional data two frequent categorical reasons for requesting telemedicine abortion were the preference to keep the abortion private (48%) and being more comfortable at home (44%). In the content analysis the wish for autonomy was a main internal motivation to choose telemedicine abortion. Women described privacy as a positive choice, the desire to choose where, when and with whom the abortion was to be performed, and the wish to choose medical over surgical abortion. One woman described this as follows.

*“I do not want to go the ‘normally’ way because my family will notice. I have extensively informed in the Internet about the pros and cons. I’m sure it’s the right decision for me. I do not want to have to discuss within the family about something that mainly concerns me.”*

#### Perception of external threat

The need to keep the abortion a secret from a family or partner was a frequent reason for requesting a telemedicine abortion in the cross-sectional study (48%) and 5% of women reported living in an abusive relationship. Adolescents were almost three times more likely to report the need to keep the abortion a secret (OR 2.78, 95% CI 1.59–4.87).

The need to keep the pregnancy and abortion a secret because of a perceived threat from the family or community was also an internal motivation in the content analysis. Over a third of women described living in a controlling environment as exemplified below,:

*“The problem is my partner. We live and work together, which means that I do not have the opportunity to go to the consultation let alone to the doctor’s practice without being noticed. (…) I am afraid he might hit me or push me against the furniture again. I would do it (the abortion) differently, but I am not able to do anything without him noticing. (transl.)”*

In the consultation form, 6% of women reported having been raped. Rape was reported with similar frequency in the email texts where it was sometimes, as exemplified below, described as exacerbating the shame associated with the abortion:

*“I was raped. I live in Germany but cannot officially have an abortion. My gynaecologist is my mother’s best friend’s daughter. If somebody sees me in a different part of town it would be a catastrophe. (…) If I disclose the rape and communicate with my family, then everything will be worse. They would not believe me. (transl.)”*

#### Shame and fear of stigma

37% of women in the cross-sectional study reported fear of stigma. Fear of stigma was further associated with living in a small town (OR = 1.47, 95% CI = 1.12–1.94) and being an adolescent (OR 1.81, 95% CI 1.08-3.01). In the content analysis, we found that self-recrimination and shame for the unwanted pregnancy were sometimes internal motivations to request telemedicine abortion. One woman described her situation as follows:

*“(…) I feel very ashamed at the thought of discussing or rather sharing my decision to abort with so many people, that I do not know. I have the feeling that one will indirectly judge me and this, in combination with the thought of having the abortion carried out by the same people, feels unbelievably difficult and wrong to me.”*

#### Financial stress

Financial hardship was a major barrier to abortion access represented with similar frequency in the cross-sectional study (40%) and the content analysis (30%). Adolescents were more likely to name cost as barrier (OR 2.61, 95% CI 1.51-4.5).

In their emails women described not being able to afford the out-of-pocket expenditure, not being able to afford to take days off work, and/or not meeting the criteria for the reimbursement of the abortion through the social welfare system. As one woman described:

*“I cannot file for reimbursement, because my income is 15 Euros above the income threshold and I have so much debt, that I can barely make ends meet each month. (transl.)”*

#### Logistic barriers

Distance as barrier to abortion care was similarly represented in the cross-sectional study (11%) and in the content analysis (10%). In their emails women often described multiple logistic barriers such as long waiting times, travel distances, work and childcare commitments and complicated bureaucracy. One woman described the complexity of the barriers facing her as follows:

*“I cannot get a doctor’s appointment. I cannot drive hundreds of kilometres, wait for weeks. I need to take care of my children, go to work. The next hospital, which does abortions, is two hours away. How am I supposed to do that? How will I get there? Where shall I leave my children during that time? (transl.)”*

#### Provider attitudes

Women described concrete prior or current negative experiences seeking abortion as their reason for choosing telemedicine abortion. These experiences included negative persuasion efforts, judgemental comments, delaying approval for abortion, or a pointed lack of assistance with the formal requirements.

One woman recounted:

*“Since I already had an abortion three years ago I know what kind of gauntlet I would have to expect. I cannot cope with this one more time.”*

#### Vulnerability of foreigners

In the emails 9% self-identified as undocumented immigrants and 21% reported difficulties related to being a foreigner. Adolescents, without specified immigration status, were more likely to report legal restrictions to access (OR 2.82, 95% CI 1.63-4.9). Self-identified undocumented residents were also highly represented among women describing financial hardship related to the abortion and controlling or threatening environments. In the content analysis the vulnerability associated to being a foreigner was a specific barrier to abortion care. These women described being denied access to services because of being undocumented or uninsured, language barriers, lack of knowledge about a complex and highly regulated system, and fear of having their immigration status revealed.

One woman stated:

*“I don’t have the proper documents, I don’t have insurance, I am an alien. Hospital won’t accept me to have a check up.””*

## DISCUSSION

This study suggests that there is a demand for alternatives to formal abortion services in Germany. Women who choose telemedicine abortion do so both from a place of empowerment, expressed as a desire for autonomy, and from a place of disempowerment, expressed as perceived barriers and fear of repercussions if the abortion were known.

The concept of access with respect to abortion is multifaceted and depends not only on legal prerequisites but on women’s attitudes, knowledge and confidence in obtaining services as well as the service delivery itself (10).

### Attitudes, knowledge, and confidence

The agency to make an informed choice is fundamental to empowerment in sexual and reproductive health and rights (SRHR) (11) (12). Women in the study who described a personal preference for telemedicine abortion had often researched their options and knew why this was a good choice for them. A previous study has shown that women who opt for an abortion in the informal sector, also in countries where abortion would be available through formal channels, often do so based on the active choice of privacy and self-management (3).

In contrast, over a third of women in this study lived in environments that limited their ability to make choices about their sexual and reproductive health (SRH) which forced them to keep the abortion a secret. Many of these women were in abusive relationships. Concealing a pregnancy or abortion from a partner is known to be associated with inter-partner violence (13).

Many women in the study also directed feeling of shame and blame about the abortion towards themselves. The systematic blaming of women in cases of sexual- or gender-based violence, or negative outcomes of pregnancy is termed “gendered blame” and is applicable also to unwanted pregnancy and abortion (9).

### Service delivery

Cost of services was a significant barrier to access in the study, consistent with a previous study from the US (17). Compared to the US, Germany has a strong public insurance system and extensive social welfare programs but abortions without medical indication are not routinely covered (18). The exemption of abortion from publicly financed health reflects a low prioritization of abortion rights.

In our results it was often the compounded effect of multiple logistic barriers that made abortion inaccessible. In a study from Great Britain barriers to abortion were suggested to result from underfunding of health services in general, impacting also abortion services (19). In Germany however waiting times to clinical appointments other than abortion are shorter than other high-income countries (20). This raises the question of specific underfunding of SRHR in Germany.

A proportion of women in the study also described negative experiences that reflect on abortion service delivery. A study in Hungary found that negative experiences with providers and fear of stigma were the main reasons women sought alternative abortion care options (21). Research supports that legalized access to abortion is difficult to enact successfully in the context of stigmatized services (22).

Undocumented immigrants in the study described formal difficulties accessing abortion, that were often exacerbated by financial hardship and lack of autonomy. Undocumented immigrants within their first 15 months of stay receive only care for acute, pregnancy- or child related health issues, excluding abortion, which makes elective interventions like abortion and family planning routinely impossible (14). Germany’s SRHR policy for immigrants resembles Switzerland’s, where undocumented immigrants show significantly higher rates of unintended pregnancies than women with documented status (15). The need for universal access to SRH has been recognized in the WHO-European Action Plan for Sexual and Reproductive Health and Rights for 2017–2021 which intends to provide a common framework for country-specific policy throughout Europe (16).

### Policy implications

This study indicates that existing services fail to provide universal access to abortion in Germany. The 1090 consultations to WoW represent over 1% of abortions performed in 2019. Abortion-related stigma persists in laws and policies and abortion care provision suffers from complicated bureaucracy and lack of information on where to access services (4, 5). The allowance of conscientious objection is known to increases the risk of judgmental treatment (18, 23). Half of Germany’s people disapprove of abortion on demand and the number of doctors who perform abortions is decreasing (24, 25).

Our results indicate that groups at particular risk of missing out on access are women with low financial means, undocumented immigrant women, and adolescents, where a particularly vulnerable group may be undocumented adolescent immigrants. Being young per say was not identified as a barrier to access in either the quantitative or qualitative data but adolescents were more likely to report lack of finances, need of secrecy, and legal restrictions compared to older women. German law requires parental consent for girls below the age of 16, which is in practice often applied to girls under 18, something which must be recognized as a barrier to access (4).

Abortion care reform in Germany requires a simplified care process, targeted interventions for vulnerable groups and sensitization of health care providers towards non-judgmental care. Even countries that formally provide elective abortion, must recognize that barriers to access in the form of scarce service delivery, stigma, prohibitive cost, or requirements of multiple appointments are enough to seriously affect women’s health and rights (26, 27).

### Strengths and limitations of the study

Motivations for women who seek abortion care outside the formal sector in Germany has not been researched. The data represented a large sample, used mixed methods which enabled triangulation, and reached information saturation.

The content analysis, based on the response to a single question, would have been nuanced by in-depth interviews. Our findings can also not make an overall assessment of abortion service delivery in Germany. A broader survey among women and providers in Germany is required to quantify the gaps in the current service delivery.

## Conclusion

Our study indicates that women in Germany who choose telemedicine abortion outside of the formal health sector do so both from a place of empowerment and a place of disempowerment. Numerous barriers exist to abortion access in the formal health sector and these may most impact vulnerable groups such as adolescents, women with low financial means, and undocumented immigrants.

## Data Availability

All data available upon reasonable request

